# Protocol for a systematic review of randomized controlled trials on nutritional interventions in individuals with obsessive-compulsive disorder

**DOI:** 10.1101/2025.07.05.25330936

**Authors:** Pau Soldevila-Matías, Joan Vicent Sánchez-Ortí, Patricia Correa-Ghisays, Vicent Balanzá-Martínez, Diego Macías Saint-Gerons, Inmaculada Fuentes-Durá, Rafael Tabarés-Seisdedos

**Affiliations:** INCLIVA - Health Research Institute, Valencia, Spain; TMAP - Evaluation Unit in Personal Autonomy, Dependency and Serious Mental Disorders, University of Valencia, Valencia, Spain; Center for Biomedical Research in Mental Health Network (CIBERSAM), Health Institute, Carlos III, Madrid, Spain; Faculty of Psychology, University of Valencia, Valencia, Spain; Teaching Unit of Psychiatry and Psychological Medicine, Department of Medicine, University of Valencia, Valencia, Spain; VALSME (VALencia Salut Mental i Estigma) Research Group, University of Valencia, Valencia, Spain; Faculty of Nursery, University of Valladolid, Valladolid, Spain

**Author notes:** These authors contributed equally to this work (Pau Soldevila-Matías, Joan Vicent Sánchez-Ortí, Patricia Correa-Ghisays). Correspondence: Vicent Balanzá-Martínez, Address: Teaching Unit of Psychiatry and Psychological Medicine, Department of Medicine, University of Valencia, Blasco Ibáñez 15, 46010, Valencia, Spain, Rafael Tabarés-Seisdedos, Address: Teaching Unit of Psychiatry and Psychological Medicine, Department of Medicine, University of Valencia, Blasco Ibáñez 15, 46010, Valencia, Spain.

**Keywords:** obsessive-compulsive disorder, nutritional supplementation, systematic review, cognitive function, quality of life

## Abstract

**Background:** Obsessive-compulsive disorder (OCD) is a chronic mental health condition, and standard treatment often results in incomplete symptom relief. Growing evidence suggests that nutritional supplements, including vitamins, minerals, and amino acids, may serve as potential adjunctive treatments for OCD by influencing neurotransmitter regulation and inflammation. Our systematic review aims to assess the comparative efficacy of nutritional supplements on relevant clinical outcomes in individuals with OCD.

**Methods/design:** A protocol was developed and registered for this systematic review. Randomized controlled trials (RCTs) assessing the efficacy of nutritional supplementation compared with standard treatments (pharmacological and psychological interventions) will be included. Primary outcomes are changes in cognitive performance, quality of life, and psychiatric symptoms. Secondary outcomes include the presence of comorbidities or multimorbidities (e.g., metabolic syndrome). Systematic searches will be conducted in MEDLINE, EMBASE, the Cochrane Central Register of Controlled Trials, and ClinicalTrials.gov from their inception onward. Two independent researchers will screen the citations and full-text articles. The risk of bias and study quality will be evaluated using validated tools, and association measures with 95% confidence intervals will be calculated. Potential sources of heterogeneity will also be analyzed.

**Discussion:** We aim to address the gaps in the current treatment paradigm for OCD by evaluating the efficacy of nutritional supplementation as an adjunctive therapy. These findings may provide insights into the potential of these supplements to improve cognitive and psychiatric outcomes. This protocol establishes a foundation for the rigorous synthesis of available evidence, which may inform future clinical practices and support the integration of nutritional strategies in OCD management.

**Systematic review registration:** Open Science Framework [https://doi.org/10.17605/OSF.IO/NZ5MU]

## INTRODUCTION

Obsessive-compulsive disorder (OCD) is a chronic and debilitating mental health condition characterized by intrusive thoughts (obsessions) and repetitive behaviors (compulsions). The lifetime prevalence of OCD is estimated to be approximately 2–3% globally, making it a significant public health concern [1]. Despite the availability of various pharmacological and psychological treatments, a significant proportion of individuals with OCD experience persistent symptoms and impaired quality of life [2]. This underscores the urgent need for innovative therapeutic approaches to improve outcomes in this population.

Recently, interest has emerged in the potential role of nutritional supplements as adjunctive treatments for OCD. Nutritional supplements, including vitamins, minerals, and amino acids, have been hypothesized to influence brain function and mental health by modulating neurotransmitter systems and reducing oxidative stress [3, 4]. For example, vitamin D regulates serotonin, a neurotransmitter implicated in OCD pathophysiology [5]. Similarly, vitamin B and glycine have been investigated for their potential to support cognitive function and reduce anxiety symptoms [6].

Existing research has examined various nutritional supplements, including vitamins D and B12 and glycine, with some studies reporting promising results in reducing obsessive-compulsive symptoms [7, 8]. Lee et al. (2019) found that vitamin D supplementation significantly improved OCD symptoms in a small cohort of patients. Kumar and Patel (2021) have demonstrated that glycine, an inhibitory neurotransmitter, reduce anxiety and improve cognitive function in individuals with OCD. However, these studies have limitations, including small sample sizes, short follow-up periods, and methodological heterogeneity. Thus, a rigorous synthesis of the available evidence is essential to draw definitive conclusions and guide clinical practice.

Moreover, the existing pharmacological treatments for OCD, primarily selective serotonin reuptake inhibitors, are associated with a range of side effects and are ineffective in all patients [9]. Psychological interventions such as cognitive-behavioral therapy are effective but require substantial time and resources, which may not be accessible to all individuals. Consequently, there is growing interest in complementary and alternative therapies, including nutritional supplementation, as additional treatment options [10].

This study aims to assess the comparative efficacy of nutritional supplements on relevant clinical outcomes in individuals with OCD. Our approach involves a systematic review of randomized controlled trials (RCTs) comparing nutritional supplements alone or in combination with standard treatments, with placebo or other active treatments. By focusing on adults diagnosed with OCD based on the Diagnostic and Statistical Manual of Mental Disorders, fifth edition (DSM-5) criteria, we aim to provide a clear and focused analysis of this specific population [11].

The specific research problem addressed in this study is the need to determine the efficacy of nutritional supplements in improving primary outcomes such as cognition, quality of life, and psychiatric symptoms, as well as secondary outcomes such as comorbidity rates [12, 13]. We hypothesized that nutritional supplementation, when combined with validated pharmacological and psychological interventions, would offer additional benefits in managing OCD symptoms compared with standard treatment alone.

## METHODS

This overview protocol has been registered in the Open Science Framework (OSF) (registration number: https://doi.org/10.17605/OSF.IO/NZ5MU). The study will be conducted according to the guidelines of the Preferred Reporting Items for Systematic Reviews and Meta-Analyses Protocols (PRISMA-P) statement [14, 15] (see **Additional File 1**). Ethical approval is not required for the conduct of this study. Data collection has been completed. Results are expected in September 2025.

### Criteria for considering studies for this systematic review

#### Types of studies

We will include RCTs that have compared a nutritional supplement alone or in combination with a standard treatment with a placebo or another alternative active treatment. Standard treatment is defined as treatment validated by the National Institute for Health and Care Excellence clinical guidelines, whether pharmacological and/or psychological.

We will exclude crossover trials because obsessive ideation and compulsions may evolve over time, and washout periods themselves may induce detrimental effects in the second period of the trial. We will also exclude cluster RCTs because of the potential for identification/recruitment bias and overestimation of the precision of the results.

#### Types of participants

We will include adults aged ≥18 years with OCD. Participants will be defined as having OCD using the DSM-5 criteria. We will exclude studies in which participants have organic causes of OCD. Additionally, we will exclude participants with comorbid psychiatric disorders such as schizophrenia, bipolar disorder, depressive disorder, eating disorders, and suicidal behavior.

#### Types of interventions

We will include nutritional supplementation (e.g., vitamin D, vitamin B12, or glycine) combined with pharmacological and psychological interventions that have been clinically validated to reduce obsessive-compulsive symptoms.

#### Comparator

The comparator will include other nutritional supplements, placebos, or an alternative active drug. Co-interventions will be acceptable in both the intervention and comparison groups provided that they are administered equally across all groups in the trial. If a trial included multiple arms, any arm that met the inclusion criteria will be included.

We will consider intervention durations of 12 weeks or longer, because nutritional supplementation is more likely to affect the relevant primary outcomes during this timeframe. We do not impose a limit on the maximum duration of intervention. Studies reporting one or more of our outcomes at least six months after the start of the intervention will be included. Therefore, the minimum follow-up duration of the studies will be set at six months.

#### Types of outcome measures

The primary and secondary outcomes are defined as outlined below. Trials will not be excluded solely because one or several of our primary or secondary outcome measures are not reported in the publication. However, if none of our primary or secondary outcomes are reported, we will include the trial only for information purposes.

I. *Primary outcomes*
  - <u>Cognition</u>: measured using validated instruments (e.g., Stroop Color and Word Test, Complutense Verbal Learning Test, Trail-Making Test)
  - <u>Quality of life</u>: measured using validated instruments (e.g., Functional Assessment Short Test and Short Form-36 Health Survey questionnaire).
  - <u>Psychiatric symptoms</u>: measured using validated instruments that include one or more of the following domains (e.g., depression, anxiety, stress, obsessions, compulsions, and psychotic symptoms, among others).
II. *Secondary outcomes*
  - <u>Morbidity or comorbidity</u>: occurrence of physical illnesses (e.g., diabetes, metabolic syndrome, or cardiovascular events) at any time before or after the initiation of the intervention with nutritional supplementation (e.g., Kaplan–Feinstein Scale, Charlson Comorbidity Index).

### Search methods for identification of studies

We will attempt to identify all relevant trials regardless of their language, publication status (published, in press, or in progress), or publication date. To minimize bias in the search results, we will develop a search strategy following the instructions in the Cochrane Handbook for Systematic Reviews of Interventions [16].

#### Electronic searches

We will search the following electronic databases: MEDLINE through PubMed (National Library of Medicine, Bethesda, Maryland, USA), EMBASE through the Elsevier platform (Elsevier B.V., Amsterdam, the Netherlands), the Cochrane Central Register of Controlled Trials (CENTRAL) via the Cochrane Library, and ClinicalTrials.gov (www.clinicaltrials.gov).

We will search these databases since their inception. The research team will design and execute a detailed search strategy using the same strategy with appropriate adjustments for each database. Search terms will include keywords related to “names/cluster of nutritional supplementation,” “cognition,” “quality of life,” “psychiatric symptoms,” “comorbidity,” and “randomized studies.” No restrictions will be imposed on the dates. The initial literature searches of MEDLINE, EMBASE, and the Cochrane Database of Systematic Reviews began in January 2025. The draft strategies for MEDLINE and EMBASE are provided in **Additional File 2**.

#### Searching other resources

We will search the reference lists of all identified trials, relevant review articles, and current treatment guidelines to identify additional potentially eligible studies.

We will contact the authors of the identified studies and research groups known to be active in pharmacological interventions for OCD to request unpublished material or further information on ongoing studies.

The search will be conducted and updated as necessary before publication. We will provide the actual dates of the electronic search during the review stage.

### Data collection and analysis

#### Selection of studies

First, two researchers will independently screen the titles and abstracts of the articles obtained from the initial searches, masked to each other’s decisions and based on the eligibility criteria. We will obtain the full texts of all potentially relevant reports. Second, these full texts will be examined in detail and screened for eligibility, with the reasons for exclusion recorded. Third, the references of all the considered articles will be manually searched to identify any relevant reports missed by the search strategy. We will examine the full articles and supplementary materials containing data and analyze their general and methodological characteristics and study results. The final versions of the articles available online will also be reviewed.

The authors will be contacted in the case of ambiguity or missing data. Duplicate records will be identified and excluded, and multiple reports from the same study will be collated such that the analysis, rather than individual reports, will serve as the unit of interest in the review. Disagreements will be resolved by consensus or consultation with a third reviewer. If disagreements cannot be resolved, we will categorize the study as “awaiting classification” and contact the authors for clarification. Studies for which relevant data cannot be obtained will also be classified as “awaiting classification.”

#### Data collection and management

Once the searches have been performed, items identified in the databases will be imported into Rayyan software (Rayyan Systems, Cambridge, Massachusetts, USA) [17]. Following the recommendations of the PRISMA 2020 declaration [18], a flow chart showing details of studies included and excluded at each stage of the selection process will be provided.

Data from each report will be used to build evidence tables describing the included studies. A standardized data extraction form will include the following information of interest: first author’s name, year of publication, type of nutritional supplementation (e.g., vitamin B12), outcomes (e.g., memory or depression), comparison, participant details (e.g., diagnostic criteria, age, sex, years of education, comorbidity, or risk factors), and maximally adjusted effect sizes, including corresponding 95% confidence intervals, hazard ratios, odds ratios, and relative risks.

### Risk of bias and quality assessment in included reviews

At least two researchers will independently assess the risk of bias and quality of each study, and any disagreements will be resolved by discussion. For the RCTs, we will evaluate the risk of bias in each included study using the Cochrane Risk of Bias 2.0 (RoB-2) [19]. This tool considers random sequence generation, allocation concealment, blinding of the participants and outcome assessors, incomplete outcome data, selective outcome reporting, and other potential sources of bias. We will judge the risk of bias for each study based on these criteria and classify the studies as having a “low,” “high,” or “unclear” risk of bias. The quality of the RCTs will also be assessed using the Cochrane Collaboration tool to assess the risk of bias [20]. This scale evaluates seven items as follows: random sequence generation, allocation concealment, blinding of participants and personnel, blinding of outcome assessment, incomplete outcome data, selective outcome reporting, and other biases [21]. Each item will be scored as “high risk,” “low risk,” or “unclear risk” based on the available evidence.

## DISCUSSION

This protocol describes a systematic review of RCTs that evaluated the efficacy of nutritional supplements as adjunctive treatments for OCD. This systematic review will establish the extent of clinical evidence supporting the association between nutritional supplementation and improvements in cognition and psychiatric symptomatology in a rigorous and reproducible manner.

Emerging evidence suggests potential benefits of nutritional supplementation as an adjunctive treatment to enhance cognitive performance, quality of life, and psychiatric symptoms in individuals diagnosed with OCD. Understanding the complex relationship between nutritional supplementation and clinical outcomes in OCD could serve as a foundation for integrating these interventions into clinical practice as complementary approaches to standard treatments. This perspective may be particularly valuable for individuals who are resistant to treatment, experience adverse effects of current pharmacotherapies, or have comorbidities.

The proposed systematic review will adhere to the reporting standards outlined in the PRISMA 2020 statement [14, 15]. Any amendments made to this protocol will be documented and outlined in the final manuscript. The results will be disseminated through presentations at scientific conferences and through publications in peer-reviewed journals. All data underlying the findings reported in the final manuscript will be deposited in the OSF (https://osf.io/), a cross-disciplinary public repository.

Finally, we aim to identify the knowledge gaps that future research can address. The implications for future studies will be discussed in the final manuscript.

The final manuscript will outline and report any amendments made to the protocol during the study. The results will be disseminated through conference presentations and publications in peer-reviewed journals. All data underlying the findings reported in the final manuscript will be deposited in a cross-disciplinary public repository such as the OSF (https://osf.io/).

## Data Availability

Deidentified research data will be made publicly available when the study is completed and published

## List of abbreviations

OCD: obsessive-compulsive disorder
RCT: randomized controlled trial
DSM-5: Diagnostic and Statistical Manual of Mental Disorders, fifth edition
OSF: Open Science Framework
PRISMA-P: Preferred Reporting Items for Systematic Reviews and Meta-Analyses Protocols
CENTRAL: Cochrane Central Register of Controlled Trials.

## Additional files

**Additional File 1:** PRISMA-P Checklist

**Additional File 2:** Key terms for MEDLINE, EMBASE, and CENTRAL search

## Declarations

### Ethical approval and consent to participate

Not applicable.

### Consent for publication

Not applicable.

### Availability of supporting data

Not applicable.

## Authors’ contributions

The study protocol was conceived by PS-M and JVS-O, with critical inputs from PC-G, VB-M, DMS-G, IF-D, and RT-S. JVS-O registered the protocol in the Open Science Framework. All the authors provided input for the study design, edited the draft protocol, and commented on the manuscript for important intellectual content. RT-S accepts full responsibility for the finished paper and organizes the decision to publish it.

## Authors’ information

PS-M: PhD (Psychology).

JVS-O: PhD (Psychology).

PC-G: PhD (Psychology).

VB-M: MD (Psychiatry),

PhD DMS-G: PhD (Pharmacy),

MSc IF-D: PhD (Psychology).

RT-S: MD (Psychiatry), PhD

---

**Additional File 1.**
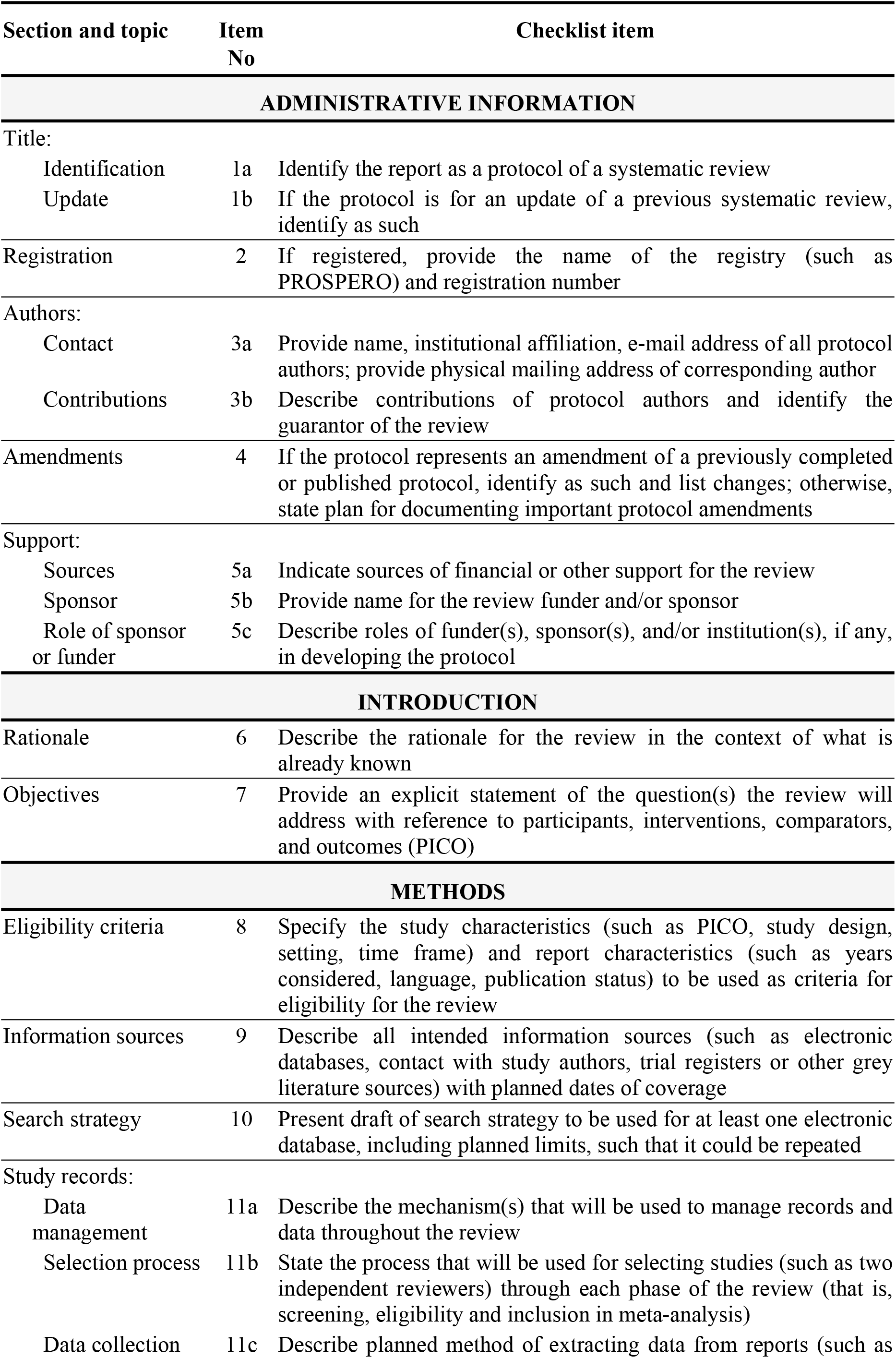
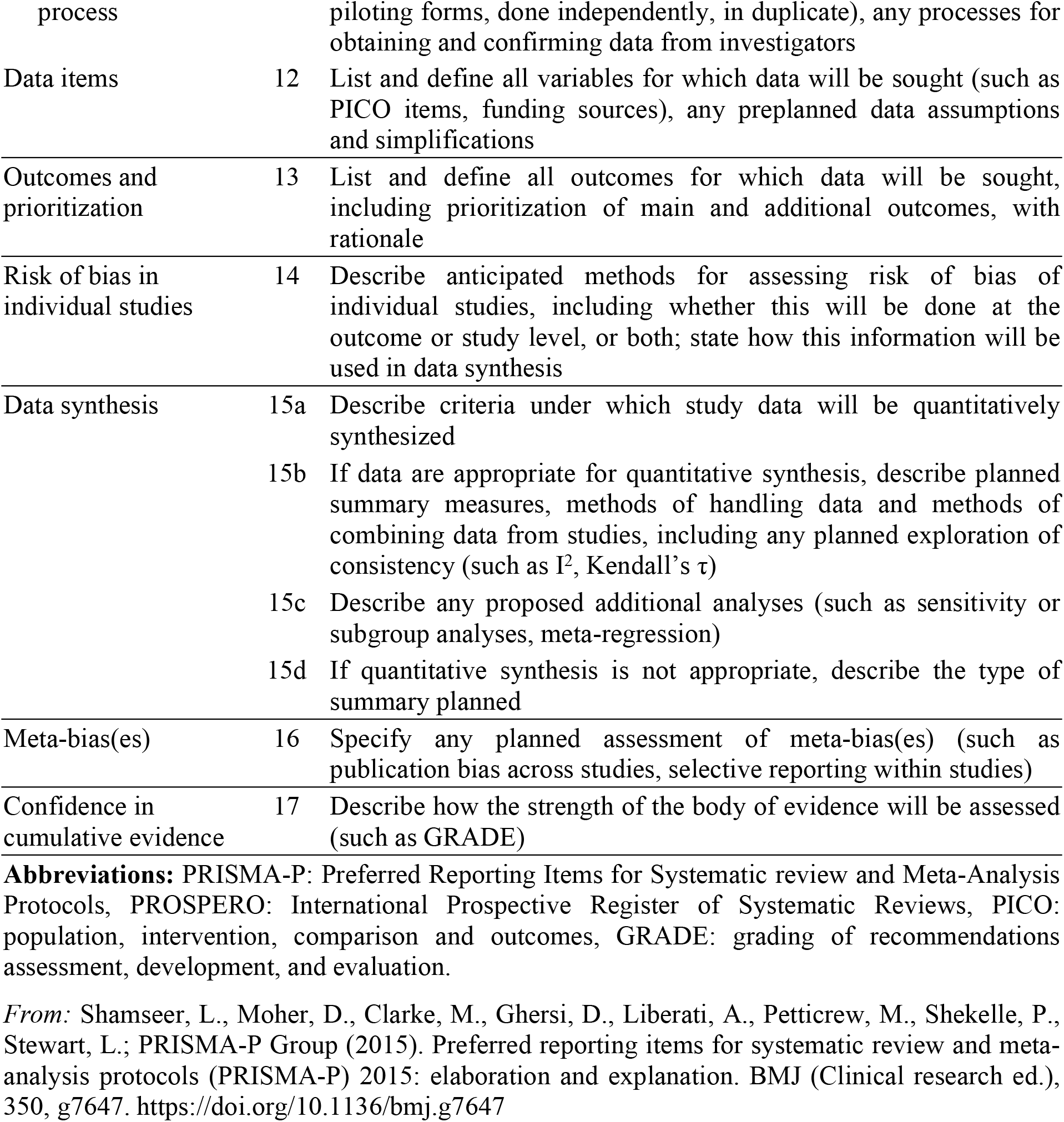
PRISMA-P checklist: recommended items to address in a systematic review protocol.

**Additional File 2.** Key terms for MEDLINE and EMBASE.

| **Search** |  | **Query** |
| --- | --- | --- |
| #1 | Diagnosis | "obsessive-compulsive disorder"[mh] OR "obsessive-compulsive disorder"[tiab] |
| #2 | Nutritional supplements | "5-HTP (5-hydroxytryptophan)"[tiab] OR "beta-carotene"[tiab] OR "biotin"[tiab] OR "calcium"[tiab] OR "choline"[tiab] OR "chromium"[tiab] OR "cobalamin"[tiab] OR "coenzyme Q10"[tiab] OR "fiber"[tiab] OR "fish oil"[tiab] OR "folate"[tiab] OR "folic acid"[tiab] OR "iron"[tiab] OR "L-arginine"[tiab] OR "L-tryptophan"[tiab] OR "magnesium"[tiab] OR "N-acetylcysteine"[tiab] OR "niacin"[tiab] OR "omega-3 fatty acids"[tiab] OR "omega-6 fatty acids"[tiab] OR "omega-7 fatty acids"[tiab] OR "niacin"[tiab] OR "niacin"[tiab] OR "pantothenic acid"[tiab] OR "potassium"[tiab] OR "probiotics"[tiab] OR "riboflavin"[tiab] OR "thiamin"[tiab] OR "vitamin A"[tiab] OR "vitamin B1"[tiab] OR "vitamin B12"[tiab] OR "vitamin B2"[tiab] OR "vitamin B3"[tiab] OR "vitamin B5"[tiab] OR "vitamin B6"[tiab] OR "vitamin B7"[tiab] OR "vitamin B9"[tiab] OR "vitamin C"[tiab] OR "vitamin D"[tiab] OR "vitamin E"[tiab] OR "vitamin K"[tiab] OR "zinc"[tiab] |
| #3 | Study type | "randomized controlled trial"[pt] OR "controlled clinical trial"[pt] OR "randomized"[tiab] OR "randomly"[tiab] OR "trial"[tiab] |
| #4 |  | #1 AND #2 AND #3 |

